# Public-private partnership to rapidly strengthen and scale COVID-19 response in Western Kenya

**DOI:** 10.1101/2021.08.31.21262891

**Authors:** Shannen van Duijn, Hellen C. Barsosio, Mevis Omollo, Emmanuel Milimo, Isdorah Akoth, Robert Aroka, Teresa de Sanctis, Alloys K’Oloo, Micah J. June, Nathalie Houben, Charlotte Wilming, Kephas Otieno, Simon Kariuki, Simon Onsongo, Albert Odhiambo, Gregory Ganda, Tobias F. Rinke de Wit

## Abstract

**INTRODUCTION:** In Africa almost half of healthcare services are delivered through private sector providers. These are often underused in national public health responses. In line with our previous HIV experience and to support and accelerate the public sector’s COVID-19 response, we initiated a public-private project (PPP) in Kisumu County, Kenya. In this manuscript we demonstrate this PPP’s performance, using COVID-19 testing as an aggregator and with semi-real time digital monitoring tools for rapid scaling of COVID-19 response.

**METHODS:** COVID-19 diagnostic testing formed the basis for a PPP between KEMRI, Department of Health Kisumu County, PharmAccess Foundation, and local faith-based and private healthcare facilities: COVID-Dx. COVID-Dx was implemented from June 01, 2020, to March 31, 2021 in Kisumu County, Kenya. Trained laboratory technologists in participating healthcare facilities collected nasopharyngeal and oropharyngeal samples from patients meeting the Kenyan MoH COVID-19 case definition. Samples were rapidly transported by motorbike and tested using RT-PCR at the central reference laboratory in KEMRI. Healthcare workers in participating facilities collected patient clinical data using a digitized MoH COVID-19 Case Identification Form. We shared aggregated results from these data via (semi-) live dashboards with all relevant stakeholders through their mobile phones. Statistical analyses were performed using Stata 16 to inform project processes.

**RESULTS:** Nine private facilities participated in the project. A detailed patient trajectory was developed from case identification to result reporting, all steps supported by a semi-real time digital dashboard. A total of 4,324 PCR tests for SARS-CoV-2 (16%) were added to the public response, identifying 425 positives. Geo-mapped and time-tagged information on incident cases was depicted on Google maps dashboards and fed back to policymakers for informed rapid decision making. Preferential COVID-19 testing was performed on health workers at risk, with 1,009 tested (43% of all County health workforce).

**CONCLUSION:** We demonstrate feasibility of rapidly increasing the public health sector response to a COVID-19 epidemic outbreak in an African setting. Our PPP intervention in Kisumu, Kenya was based on a joint testing strategy and demonstrated that semi-real time digitalization of patient trajectories in the healthcare system can gain significant efficiencies, linking public and private healthcare efforts, increasing transparency, support better quality health services and informing policy makers to target interventions. This PPP has since scaled to 33 facilities in Kisumu and subsequently to 84 sites in 14 western Kenyan Counties.

## Introduction

Most health systems in low and middle income countries (LMIC) are underfunded and understaffed, with limited isolation and intensive care infrastructure^1,2^. In sub-Saharan Africa, health systems are facing disproportional challenges and by definition are ill-equipped and under-resourced to deal with additional burdens, such as those caused by the recent COVID-19 pandemic. There is an urgent need for innovative approaches to accelerate strengthening African health systems to work towards Universal Health Coverage (UHC).

Kenya confirmed the first COVID-19 case on March 13, 2020. As of November 28, 2021, 254,951 confirmed cases with 5,333 fatalities had been reported^3^. By mid-March 2021, Kenya recorded the beginning of its third wave with a notable steep increase in daily COVID-19 cases and deaths^4,5^. This third wave led to stringent measures, particularly in the capital city Nairobi and nearby Counties (Kajiado, Machakos, Kiambu and Nakuru)^6^. The third and fourth wave were with Gamma and Delta variants, while the fifth one at the end of 2021, early 2022 was with Omicron^7^. During summer 2022 a sixth wave is evolving (Omicron BA4, BA5).

On March 4 2021 Kenya received the first batch of COVAX COVID-19 AstraZeneca vaccines, prioritizing vaccination of its high-risk population, including frontline healthcare workers, adults above 58 years, teachers, police officers and persons with pre-existing conditions^8^. However, the first weeks of roll-out were met with considerable vaccine hesitancy amongst various target groups, which lasts until today^9^. Kenya aimed to vaccinate 30% (15 Million) of its total population of 50M by the end of June 2023^10^. This vaccination target falls significantly below the 70% goals set for the world by WHO and for Africa by CDC-Africa^11,12^.

In the absence of vaccine-induced immunity, the options to combat COVID-19 in Kenya were relatively limited and mostly include so called non-pharmaceutical interventions, including lockdowns, curfews, social distancing, personal hygiene and protective clothing, particularly at healthcare facilities. When diagnostic testing became available, those who appeared SARS-CoV-2 infected were (advised to) quarantine(d). At the start of the pandemic, diagnostic testing for SARS-CoV-2 was led by the Kenyan MoH at the national and county levels through centralized PCR testing in dedicated high-throughput laboratories, with KEMRI playing a central role. However, centralized testing in Kenya faced multiple challenges, including lack of funding, stockouts of reagents and testing kits, manpower challenges, PCR equipment breakdowns due to heavy workload resulting in test access limitations and prolonged test turnaround time. During most of 2020, RDTs for SARS-CoV-2 according to WHO standards were unavailable in Kenya. In December 2020 the first Kenyan interim guide for RDTs was launched^13^. MoH supported COVID-19 testing remained confined to designated public hospitals with limited tapping of the private sector’s potential^14^.

The private sector (for-profit, not-for-profit foundations and faith-based organizations) is a significant player in health service delivery in sub-Saharan Africa^15^ and particularly in Kenya^16^. The private healthcare sector can add substantial capacity to the public health infrastructure, which often faces challenges in terms of quality of care, drug stockouts, health worker shortages, industrial action, and lack of diagnostic equipment^17^. PPPs can play an essential role in LMICs health system strengthening^9^, particularly during outbreaks and epidemics, where a coordinated, rapidly scalable approach is required. Strengthening and coordination of public and private health systems is needed to ensure progress towards UHC and global health security^18^. The challenge is to combine private and public efforts in healthcare delivery in a mutually supportive and collaborative manner. Achieving a supportive PPP is complex, difficult to form, fraught with challenges and evidence of their effectiveness is limited^19,20^. There are ample examples of (inter)national responses where private healthcare sector initiatives were crowded out by parallel public sector efforts^21^. Crowding out implies that private investments in healthcare are replaced instead of supplemented by public funds, and the total amount of funds in the healthcare system remains unchanged. If supported well, PPP models can enhance capacity, increase quality of services offered, promote access, and offer innovative and sustainable solutions to healthcare challenges in developing countries^22^.

The Dutch NGO PharmAccess has gained extensive experience supporting innovative PPP models for healthcare, ‘crowding in’ private funding. Notable PPP models include the first risk equalization fund for HIV in Africa^23^, the first Medical Credit Fund for Africa that provides loans to private-sector health entrepreneurs through public-private funding^24^ and digital technologies provided through the M-TIBA platform to support Kisumu County’s UHC^25^. Due to PharmAccess’ experience with timely interventions and through rapid donor contributions and necessary regulatory support by the local Department of Health, a unique joint PPP named “COVID-Dx” was started in early May 2020 in Kisumu County, Kenya. COVID-Dx was designed to enhance Kisumu’s capacity for COVID-19 sample collection and testing, rapid digital clinical and socio-demographic data collection and timely reporting to inform policy decisions. The COVID-Dx intervention connected private- and faith-based healthcare facilities in Kisumu County to the existing MoH network, it implemented COVID-19 clinical guidelines, it enabled COVID-19 testing at the

KEMRI central laboratories and it created semi-real time dashboards to improve reporting efficiency and decision-making by both healthcare workers and policymakers. The aim of this study is to describe the performance of a rapid PPP in responding to the COVID-19 pandemic, in a sub-Saharan African setting and to underscore the importance of digitalization of the health system to do so. Determinants of acceptability by health workers are described elsewhere^26^.

Kisumu County, located in the western part of Kenya, was selected for this PPP because of its unique track record as a county pioneering UHC using the M-TIBA digital health platform^25^. This creates the potential of reaching out to the entire 1.2M population and work on a replicable model of digital infrastructure that can serve future epidemic preparedness for settings in Kenya. The intervention was well-timed, starting exactly at the time that Kisumu reported the first two COVID-19 cases on May 27, 2020. As of August 25, 2021, Kisumu had reported 8,897 cases and 285 deaths out of a total of 48,006 PCR tests and 15,409 rapid antigen tests done^27^.

## Material and Methods

### Context

After a preparation phase, COVID-Dx officially started in Kisumu, Kenya on June 1, 2020, and its first phase, that this paper reports about, ended March 31, 2021. Healthcare facilities in Kisumu were selected for participation based on a set of criteria, including: possession of a valid license, a MoH COVID-19 certificate, being within reasonable geographic distance from KEMRI testing laboratories, serving minimally 100 patients per week, participating in the PharmAccess SafeCare quality improvement program, being connected to M-TIBA^28^, having an average staff of at least 25, possessing an operational and regularly serviced fridge and generator for sample storage and proving higher management willingness to participate in COVID-Dx. Nine healthcare facilities were eligible, labelled as A-I throughout this manuscript to ensure anonymity. Table 1 provides an overview of the key characteristics of these healthcare facilities. Facility A was a small facility used a pilot to test steps of the COVID-Dx intervention before scaling to the other 8 facilities. Each participating facility had trained staff collecting nasopharyngeal and oropharyngeal swabs from patients who fulfilled the COVID-19 case definition as per the Kenyan Ministry of Health COVID-19 Clinical Case Definition Guidelines. The main eligibility criteria to be tested for COVID-19 were: 1) people presenting with signs and symptoms of COVID-19, fulfilling the criteria of the so called COVID Clinical Identification Form (CCIF) and 2) risk groups as defined in the Kenyan MoH guidelines: healthcare workers, contacts of confirmed COVID-19 cases, travellers from high-risk areas^29^. Participation of patients was completely voluntary.

**Table 1:** Overview of key characteristics participating COVID-Dx facilities.

| Facility | Kenyan Healthcare category | Patients/day | Beds | SafeCare level <sup>30</sup> | Staff | Setting | Start project | First case identified |
| --- | --- | --- | --- | --- | --- | --- | --- | --- |
| A | Level 3 Health Centre | 40 | 0 | 3 | 12 | Urban | Jun 2020 | Aug 2020 |
| B | Level 4 Hospital | 100 | 100 | 2 | 80 | Urban | Jul 2020 | Sep 2020 |
| C | Level 4 Hospital | 100 | 60 | 3 | 60 | Rural | Jul 2020 | Sep 2020 |
| D | Level 5 Hospital | 180 | 86 | NA | 200 | Urban | Aug 2020 | Aug 2020 |
| E | Level 4 Hospital | 70 | 62 | 3 | 50 | Urban | Aug 2020 | Sep 2020 |
| F | Level 5 Hospital | 50 | 50 | NA | 80 | Urban | Sep 2020 | Sep 2020 |
| **G** | Level 5 Hospital | 300 | 180 | NA | 400 | Urban | Dec 2020 | Dec 2020 |
| **H** | Level 5 Hospital | 100 | 70 | 5 | 400 | Urban | Dec 2020 | Jan 2020 |
| **I** | Level 5 Hospital | 1000 | 550 | NA | 600 | Urban | Feb 2021 | Feb 2021 |

### Laboratory methods for SARS-CoV-2

Patient samples were collected in viral transport medium according to manufacturer’s instructions (F&S scientific^31^) and transported by motorbike in cool boxes to the KEMRI central laboratory for SARS-CoV-2 RT-PCR testing within 24 hours. Results were reported through KEMRI and the MoH system to participating providers who reported to patients usually within 24–48-hours. Complementary (telephone and personal) counseling services were provided involving two counselling session per client (pre and posttest counselling). The duration of the phone calls could range from less than 5 minutes to more than 10 minutes per client, depending on the situation at hand. KEMRI central laboratory trained staff carried out the PCR test procedures according to standard manufacturer prescribed testing protocols. Laboratory staff used MagMAX™ Viral RNA Isolation Kit^32^ to manually extract SARS-CoV-2 viral RNA from the paired nasopharyngeal and oropharyngeal samples. Post-RNA extraction, the TaqPath™ 19 kit^32^ was used to carry out real-time SARS-CoV-2 PCR. Laboratory staff employed the following thermocycling conditions; 2 min at 25°C incubation, 10 min at 53°C for reverse transcription, 2 min at 95 °C for enzyme activation and 40 cycles of 3 s at 95°C and 30s at 60°C. Samples having exponential growth curve and Ct < 40 in at least two SARS-CoV-2 targets were considered positive. Between December 28, 2020, and March 31, 2021, an additional prospective diagnostic evaluation of a rapid antigen kit was carried out and the results of this evaluation are reported in a separate paper^33^. All Ag-RDTs were followed by a confirmatory PCR test, which is the basis for the quantitative analyses reported in the current manuscript.

### Use of Digital Tools

The official Kenyan CCIF used by the MoH to screen and report all COVID-19 cases was digitalized to run on simple tablets that were distributed to participating providers. The digital CCIF tool additionally collected important logistical information including full tracking and tracing of samples and data flows: sample collection, courier receipt, road transport, KEMRI laboratory receipt and triaging into various sub-laboratories, specifics of PCR tests performed, result verification, transmission of final diagnostic result to MoH and finally to healthcare provider for release to patients reporting. The CCIF-Tool data was stored in a dedicated database (CommCare; Dimagi), a robust mobile data collection and service delivery platform hosted in a highly secured ISO27001 environment that is HIPAA and GDPR compliant. KEMRI provided oversight of scientific accuracy and quality of SARS-CoV-2 testing data. Aggregated results from the CommCare database were shared via a PowerBI dashboard with pertinent policymakers and relevant stakeholders through password-protected personal tablets and mobile phones. Microsoft PowerBI is a collection of software services, apps and connectors that visualize interactive insights of data sources^34^. The dashboard presents an overview of the clinical and socio-demographic characteristics of patients tested, positivity rates, participating facilities, and maps with the patients per place of residence. The supplementary materials include a screenshot of the dashboard (Supplementary Figure 1).

### Data analyses

We conducted data analyses for operational purposes to inform and improve project processes, monitor general progress of the project, inform participating providers and generate aggregate data to inform policy makers. We performed descriptive statistical analyses using Stata 16, tested relationships between categorical variables using chi-square tests, and used an independent sample t-test to explore relationships between categorical and continuous variables. We considered a p-value of <0.05 to be statistically significant during the analyses.

### Ethical clearance

Ethical clearance for this project was obtained from Jaramogi Oginga Odinga Teaching & Referral Hospital (JOOTRH) on June 16, 2020, with approval number IERC/JOOTRH/230/2020. KEMRI also provided ethical clearance on September 30, 2020, with approval number KEMRI/RES/7/3/1. Research License was obtained from the National Commission for Science, Technology, and Innovation (NACOSTI) on July 6, 2020 (NACOSTI/P/20/5616).

### Patient and Public Involvement

The Kisumu DoH, KEMRI and participating healthcare facilities were actively involved in co-creating the design, conduct, reporting, and dissemination plans of the research. Feedbacks from patients and public was collected through ongoing counselling services and fed back into the COVID-Dx operations to further improve.

### Building the PPP model

In Table 2, we present the main tasks that needed to be executed during the PPP and which of the PPP stakeholders (gradually) took the lead in executing that task. Choices regarding task division were based on the partners’ different levels of authority, partner’s capabilities, and capacities, such as time, knowledge about regulatory or laboratory processes, and staff available. In the case of this PPP, the stakeholders were all operational on different levels, and authority levels of each partner were clear for the other partners. If unclarities emerged, these were resolved by weekly stakeholder meetings coordinated by PharmAccess. Generally, the DoH was responsible for approving selected participating facilities, providing guidelines, performing trainings, carry out contact tracing, and application of epidemic control guidelines. PharmAccess provided kickstart funding, managed day-to-day operations, supported psychosocial counselling to clients and providers staff, procurement of reagents and personal protective equipment (PPE), database hosting, cleaning, quality control and translation into the COVID-Dx data dashboard. KEMRI provided supportive supervision, complementary training, conducted social research and was responsible for the full laboratory component (SARS-CoV-2 testing and RDT evaluation). Healthcare facilities executed patient management, sample taking, data entry into the CCIF-tool, assisted with contact tracing of COVID-19 patients, disseminated test results to patients and provided counselling.

**Table 2:**
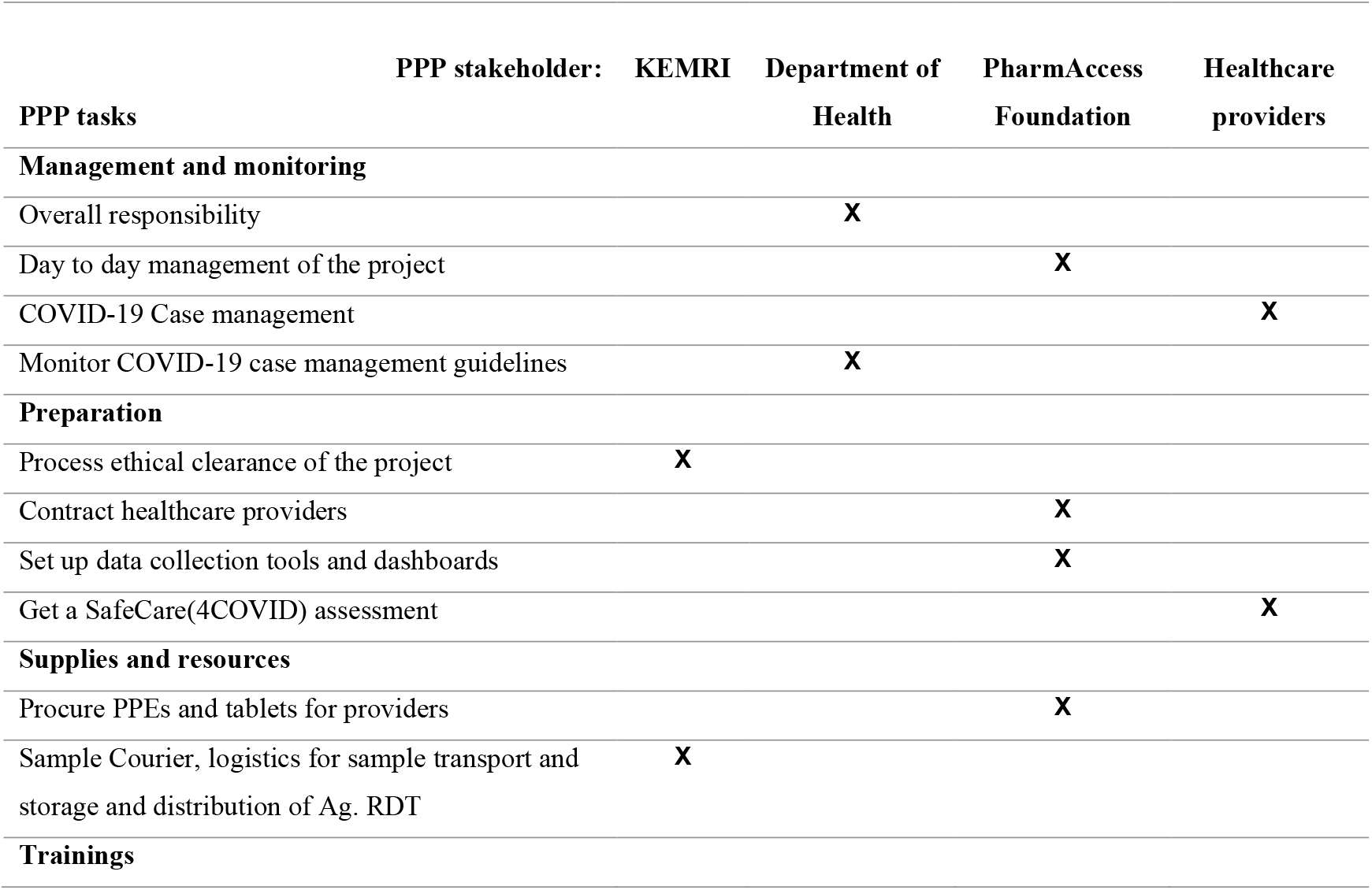
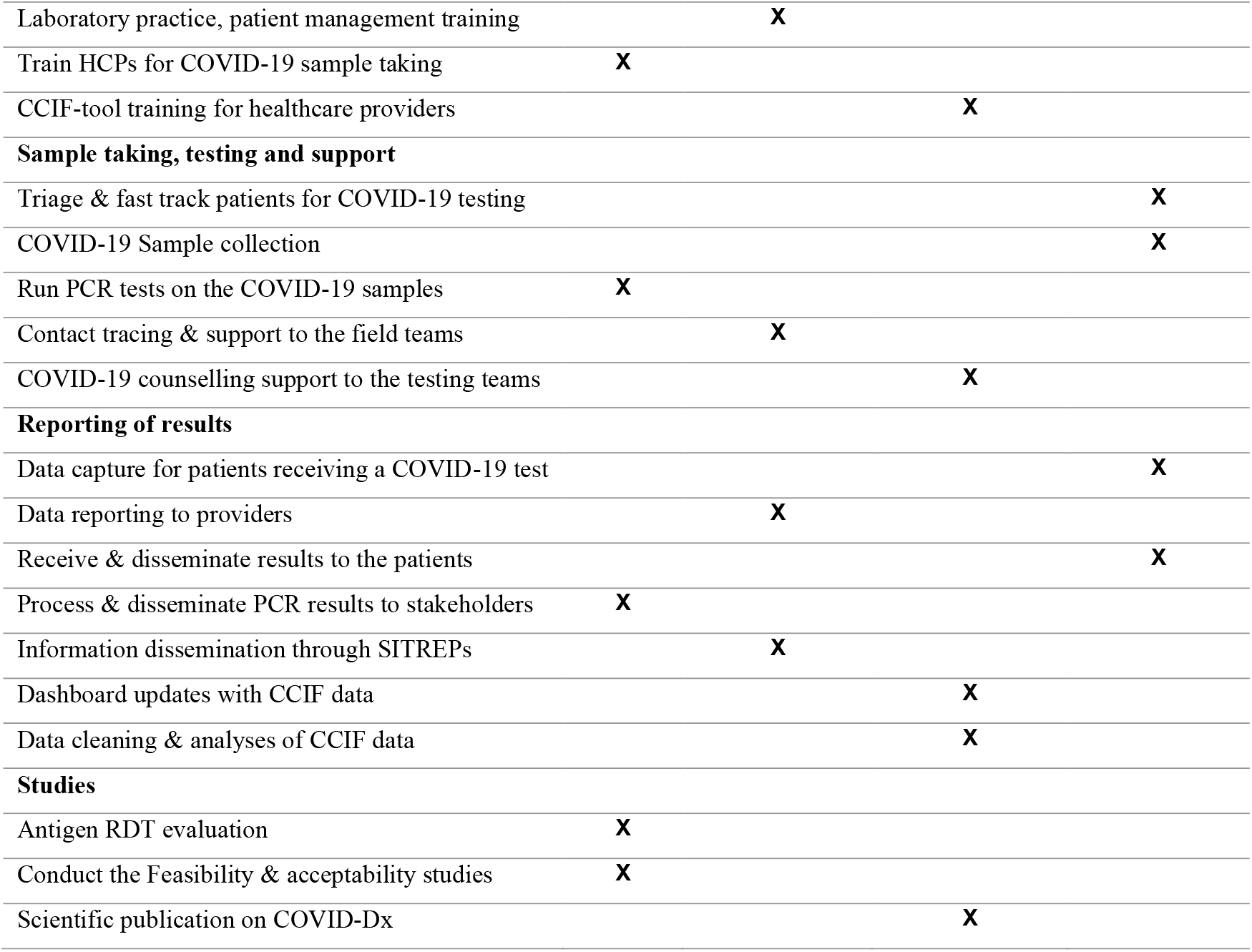
Roles and tasks of public and private partners in COVID-Dx project. *for each task the leading institution is indicated.

## Results

### Key events

The supplementary materials include a Gantt Chart (Supplementary Figure 2), outlining our key experiences during this PPP. It can be seen that preparations for COVID-19 were starting well before Kisumu County reported the first COVID-19 cases on May 27, 2020. At the onset of COVID-Dx, patient sample collection and central testing via MoH-linked KEMRI laboratories was restricted to designated public hospitals. MoH supported a limited set of public hospitals collecting samples and central laboratories testing for COVID-19 with providing PPE, sample collection materials, sample transportation materials such as viral transport medium (VTM), cool boxes, and sample testing reagents.

For private facilities to be added to this initial COVID-19 response and have access to public sector COVID-19 testing services through the KEMRI central laboratories, it was initially indicated that COVID-Dx had to be positioned as a research project. This implied a phase of comprehensive protocol writing and ethical clearance procedures. Later it was clarified that any private facility intending to provide COVID-19 testing through MoH-supported central laboratories were required to do so via the MoH COVID-19 public sector response at the County level. This meant selected private healthcare facilities just required DoH approval as official sample collection sites supporting Kisumu County to scale up COVID-19 testing.

In the PPP preparation phase, multiple meetings were held with various players to align project objectives culminating in signed contracts with expected deliverables clearly outlined. Selected private providers as approved by the DoH were subsequently contracted. These private providers gradually became known to the public as additional COVID-19 service sites featuring free diagnostic tests. In the first months of implementation multiple trainings were performed for providers (COVID-19 Guidelines, safety measures, sample collection, sample transport, data entry using tablets) in full coordination with the Kisumu DoH that provided training of trainers. During COVID-Dx roll-out regular complementary refresher trainings were organized by PharmAccess.

Several procurement rounds of PPEs occurred during the project (May, August, October, December 2020). At project initiation, the PowerBI digital dashboard was developed to share the live operational results of the project. The dashboard was continuously improved and extended throughout the project, based on continuous feedbacks of the users. In addition to the healthcare providers, by November 2020, the first external stakeholders (policy makers) got access to the dashboard.

Some months into the project, several healthcare facilities noticed fewer patients coming in to get tested. Increasing COVID-19 stigma (due to widely feared quarantine measures) turned out to be associated with hesitancy in testing and avoidance of healthcare facilities. Additionally, Kisumu County DoH mandated patient contact tracing. As the positive cases increased, COVID-Dx provided complementary contact tracing services to the DoH. Moreover, PharmAccess contributed a senior counsellor to COVID-Dx, who provided mental support to clients and in addition trained the participating facility-based counsellors to address COVID-19 stigma. Most facility-based counsellors were already HIV counsellors, with previous experience, which facilitated the COVID-19 counselling training.

At times the DoH or KEMRI requested some private facilities to stop testing or the federal MoH restricted the criteria that selected patients for testing. For instance, around October 2020, KEMRI and DoH temporarily paused sample collection from one of the COVID-Dx facilities when samples spilt during transportation, creating a health and safety hazard. This incident triggered an audit and mandatory refresher training of the affected site. Also, the MoH updated guidelines for targeted testing, where only those who met the CCIF case definition and had symptoms were eligible for testing; this continued for several months, restricting testing to fewer patients. Progress of the project was disseminated regularly through stakeholder involvement meetings, supplemented with the semi-real-time mobile phone dashboard.

## Quantitative data

### Project impact

Through nine participating facilities, the COVID-Dx project supported a total of 4,324 PCR tests for SARS-CoV-2, of which 425 tested positive (194/2,138 female and 231/2,186 male). There was no significant association between gender and test result, X^2^(1, *N*=4,324) =2.6, *P* =0.11. The overall COVID-19 positivity rate during the project was 9.8%. Figure 1 presents weekly positivity rate based on PCR tests during project implementation. As shown, there were no COVID-19 cases found during the first months of the project (June 2020 and July 2020). We noted a peak in COVID-19 cases in December 2020, which mirrored the Kenyan ‘second wave’, with a positivity rate of 23.1% at its highest point.

**Figure 1.**
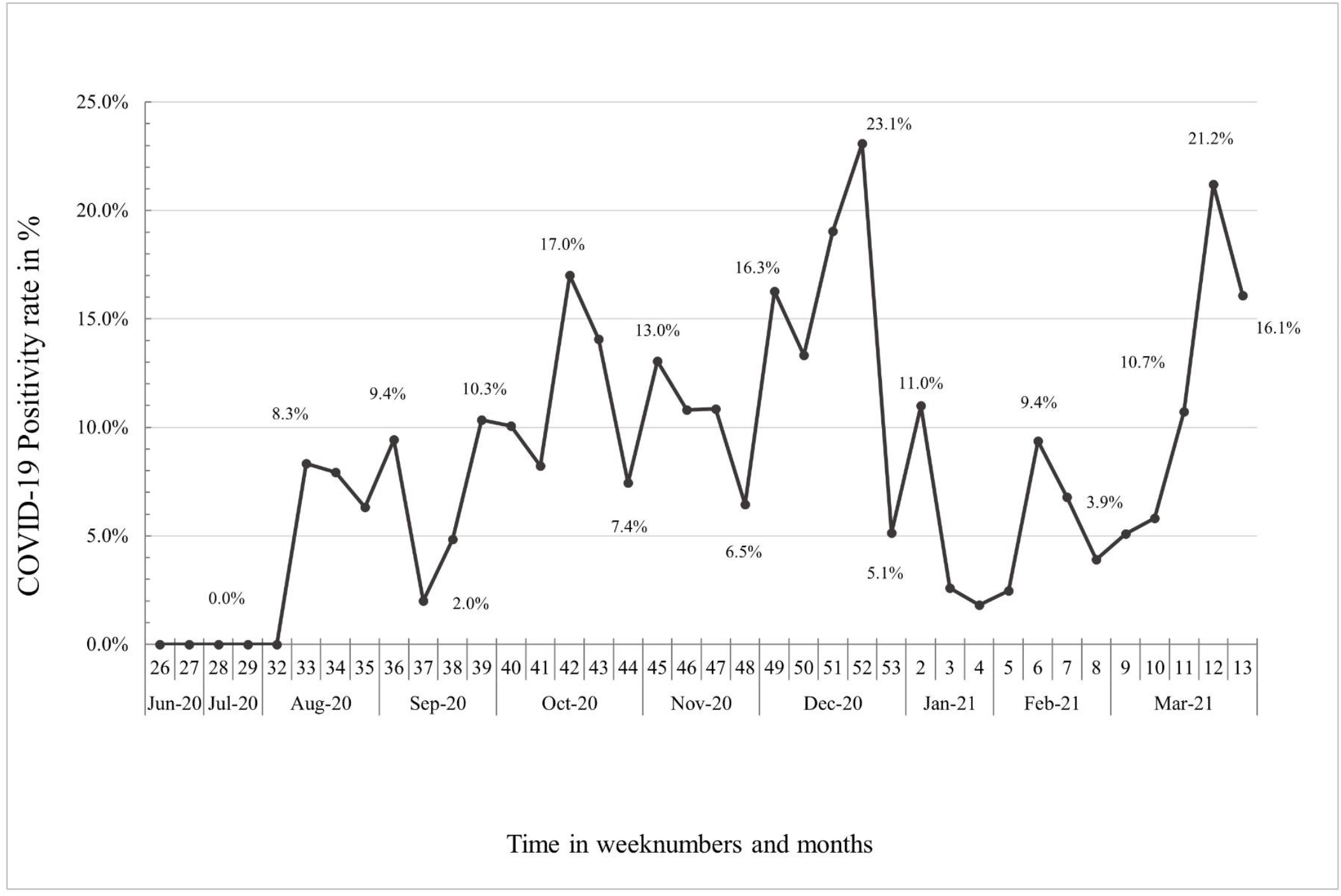
PCR positivity rate over time in COVID-Dx project; June 2020 - March 2021.

Table 3. provides an overview of COVID-19 positivity rates per participating facility. Positivity rates of facilities varied between 31.9% (facility H) and 3.8% (facility B). Figure 2 shows the percentage of positive cases per facility over time. In this graph, we show that in November 2020, there was an overall low point in positivity rate for all facilities. In February and March 2021, the positivity rate rose to its then highest point.

**Table 3.** COVID-19 positivity rates per healthcare facility.

| Facilities | Positive cases | Total tested | Positivity rate |
| --- | --- | --- | --- |
| A | 17 | 348 | 4.9% |
| B | 11 | 286 | 3.8% |
| C | 52 | 1023 | 5.1% |
| D | 130 | 598 | 21.7% |
| E | 7 | 41 | 17.1% |
| F | 25 | 224 | 11.2% |
| G | 93 | 948 | 9.8% |
| H | 53 | 166 | 31.9% |
| I | 37 | 662 | 5.6% |

**Figure 2.**
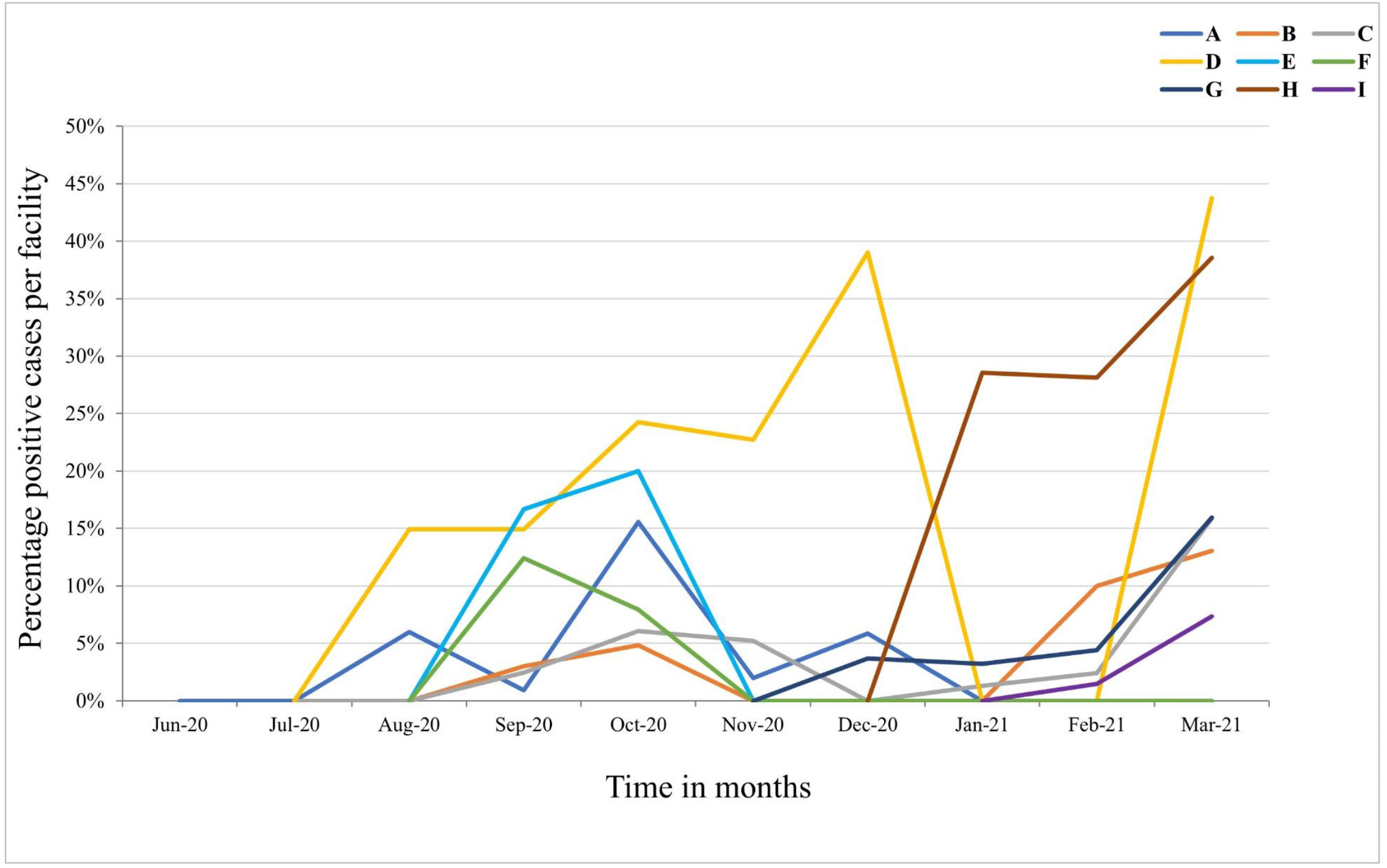
Percentage of CO VID-19 cases per facility per month; June 2020 - March 2021

### Clinical presentation

Patient reported clinical symptoms were captured using the CCIF-Tool. Reported symptoms from COVID-19 positive patients in order of frequency included: cough, headache, general weakness, and history of fever/chills. Figure 3a. presents an overview of the most common symptoms reported for all cases and COVID-19 positive cases. Of note, 22.8% of all positively tested patients were asymptomatic. Twenty-two percent of positive cases reported pre-existing conditions, with cardiovascular disease as the most common; 9.9% of all positive cases had cardiovascular diseases (n=42 times), and 5.6% (n=24) had diabetes. Figure 3b. presents the percentage of asymptomatic cases which tested positive per age group. This figure demonstrates a significant inverse correlation between age and COVID-19 positivity (X^2^(6, *N*=432) =27.4, *P* =0.00). Figure 4. shows the number of tests performed per age group during the project for all participating facilities. The average age of negative tested patients was 36.4 years, and the average age of positive tested patients was significantly higher at 40.2 years (two-sample t-test (M= 36.8, SD = 15.4), 95% CI [36.3, 37.3], t = -4.8, *P* = 0.00).

**Figure 3a.**
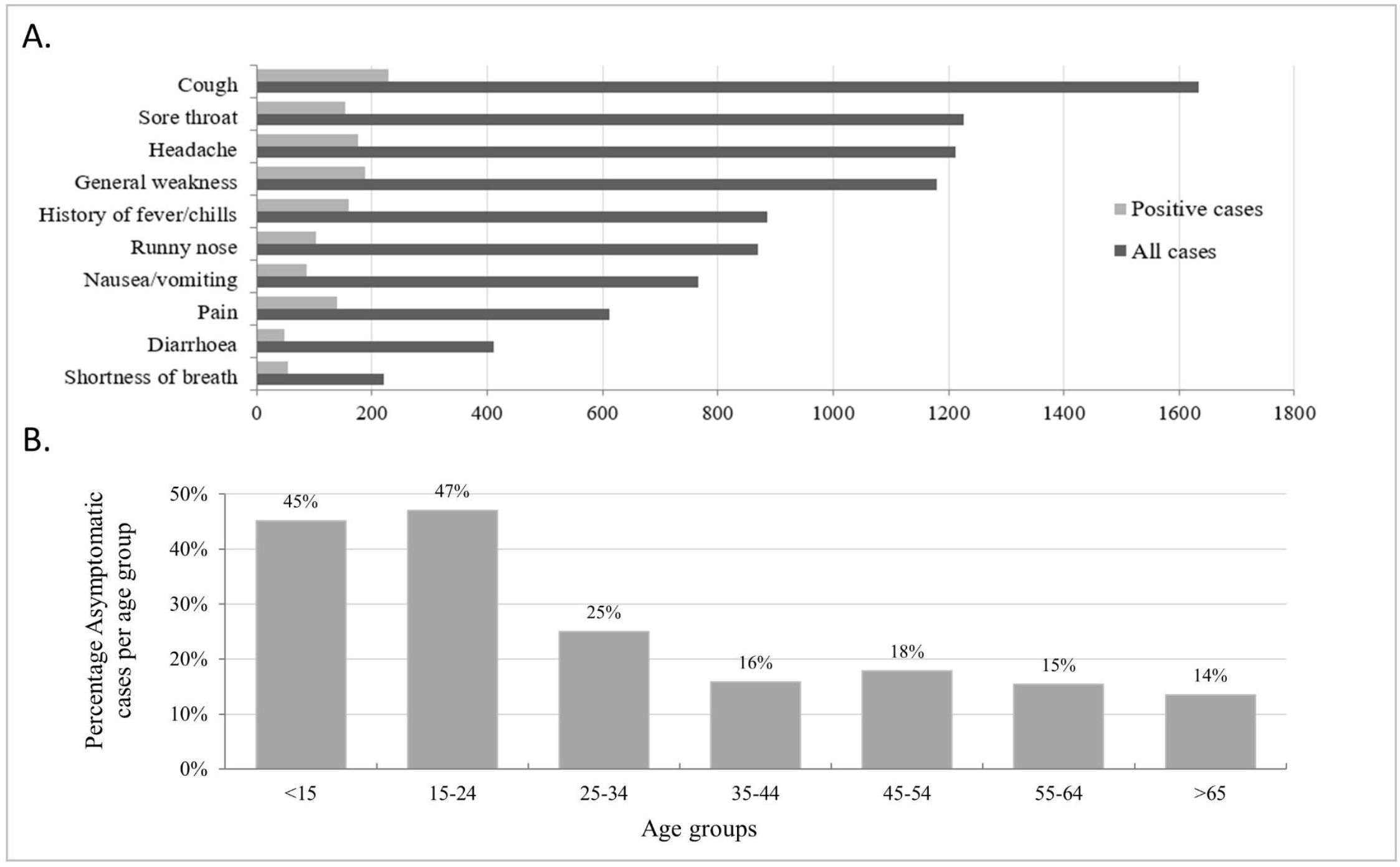
Overview of most common symptoms reported during COVID-Dx project roll-out (all cases versus COVID-19 cases). **Figure 3b**. Percentage asymptomatic COVID-19 patients per age group

**Figure 4.**
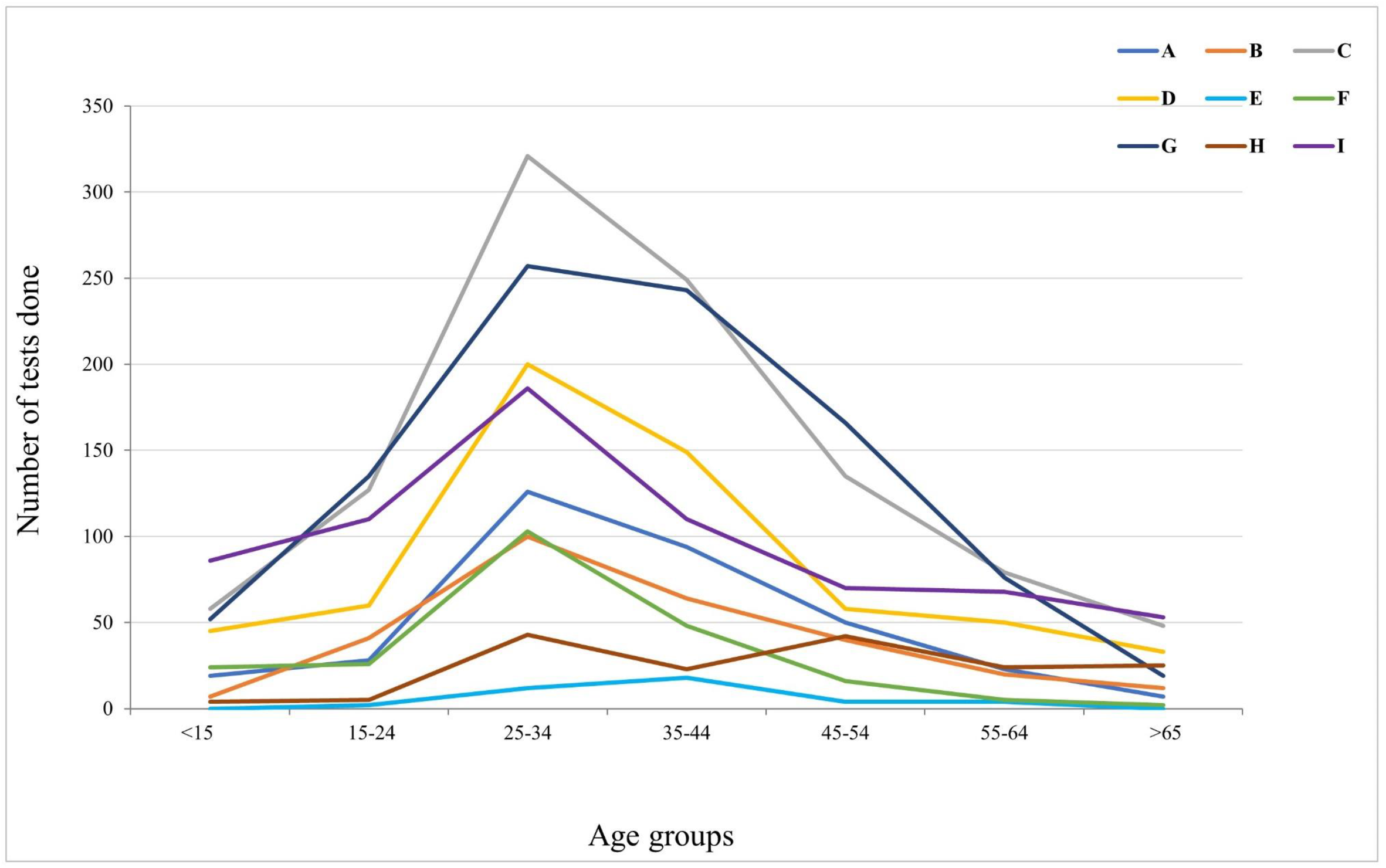
Number of SARS-CoV-2 tests done per age group per facility

### Healthcare workers

At these early stages of the COVID-19 epidemic, our PPP particularly prioritized healthcare workers who were at increased risk at the frontline of healthcare delivery. We performed 1,009 SARS-CoV-2 tests on healthcare workers (23.3% of total), representing 43% of total Kisumu County healthcare workforce. We noted a positivity rate of 7.6% (n=77) among healthcare workers, which was slightly but significantly lower than the overall (9.8%) positivity rate (X^2^(1, *N*=4324) =7.5, *P* = 0.01). Moreover, 36% of tested healthcare workers who tested positive were asymptomatic. Other regularly tested groups were the self-employed (12.5%, n=539) and students (11.0%, n=476).

### Scalability

In early Q2 2021 when this PPP was virtually coming to an end, a sudden outbreak of COVID-19 Delta variant took place at a sugar estate in Kisumu^35^. This resulted in local panic, given the well-known images of the consequences of infection with this variant in India. The DoH requested immediate support from COVID-Dx, which was provided in the form of procurement of RDTs for ring-fencing. More importantly, the COVID-Dx PPP team developed within 48 hours a new dashboard that could accommodate data from 33 public and private health facilities in Kisumu. Up to date this dashboard has accumulated almost 50K test results, ten-fold the original number realized by the 9 private facilities (Figure 5). Later in 2021 (Q3), the Lake Region Economic Bloc, a consortium of 14 Counties in West Kenya covering a population of 15 million Kenyans expressed its interest to adopt COVID-Dx. This subsequently was realized, with 84 participating COVID-19 service points and ∼60K tests performed up to today. The latest development is the transition of COVID-Dx into Epi-Dx, a digital platform that not only covers COVID, but also 19 additional epidemical diseases corresponding to the official Kenyan IDSR tool^36^. Again, rapid scaling was quickly realized due to the digital framework of COVID-Dx and the trust already established working with PPP stakeholders.

**Figure 5.**
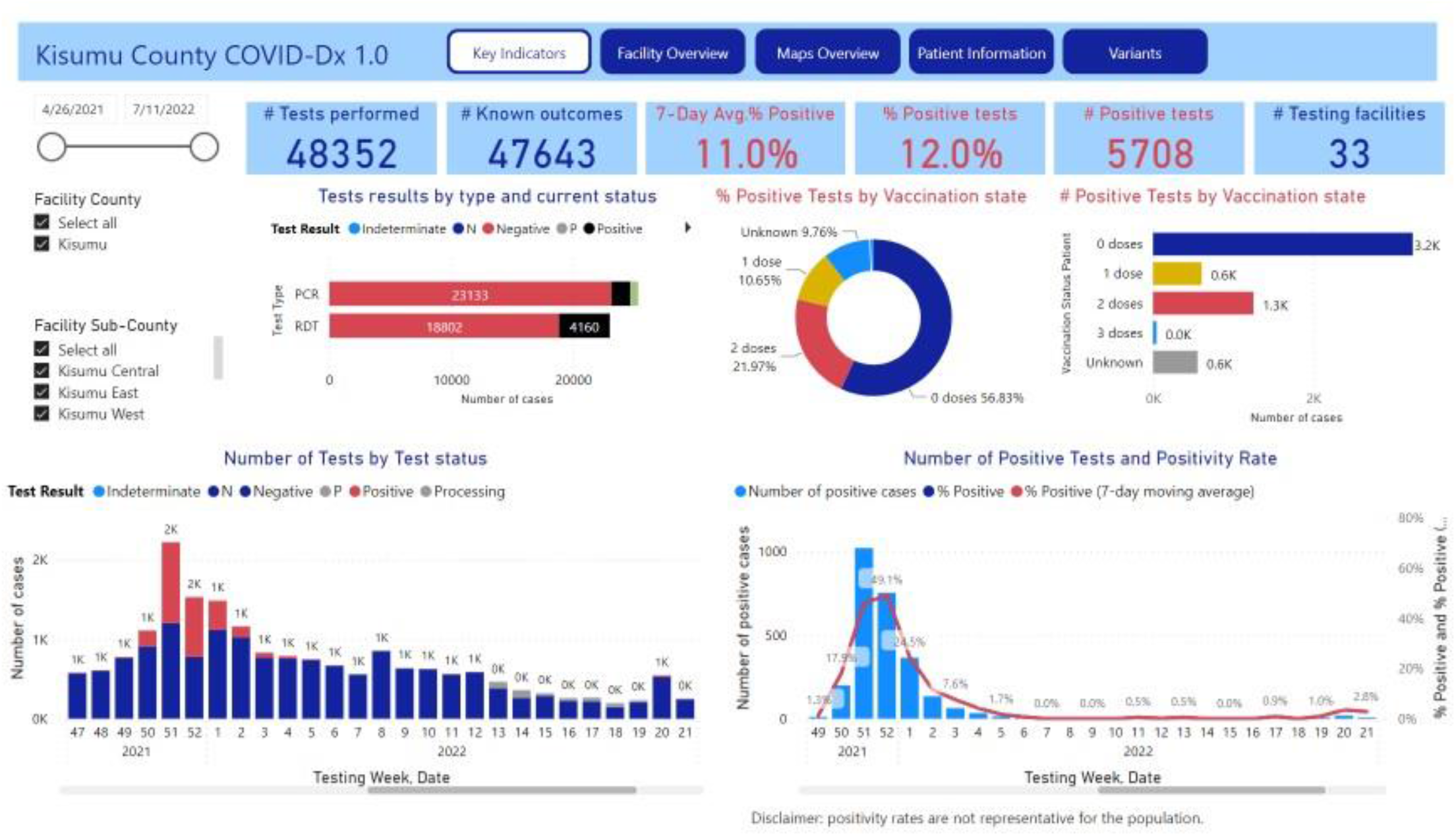
Screenshot Dashboard COVID-Dx as per July 2022

## Discussion

### Qualitative insights

This paper describes experiences and findings during a rapid PPP intervention responding to the COVID-19 pandemic in Kisumu, Kenya. Lessons learnt appeared scalable and could serve epidemic preparedness, in comparable sub-Saharan African settings. Typically, COVID-19 responses in Africa are addressed through ‘vertical’ programs rolled out mainly through the public sector. This was done for HIV in the past, leading to parallel healthcare delivery and financing infrastructure in Africa and other LMICs, that is crucially dependent on international funding mechanisms through institutions like the GFATM and PEPFAR.

Today, in the UHC era ways should be explored that ‘horizontalize’ vertical disease-specific funds into primary healthcare delivery. This paper presents such a model: integrating private healthcare facilities into the public COVID-19 response and supporting this with a full digital trajectory tool. In Kenya, almost half of healthcare is delivered through private healthcare facilities. Therefore, at the onset of COVID-19, with then-unknown consequences, we opted for an immediate scaling of healthcare capacity to deliver COVID-19 services by creating a PPP model. This was informed by our experience with HIV service delivery starting in the private sector in Africa and expanding to the public sector^37^. In addition, we wished to make use of important digital developments that are taking place in Africa and particularly Kenya that include the mobile phone revolution, the development of bankless banking through M-PESA, the digitalization of health data exchange (M-TIBA), the availability of smart-phone apps to support data dashboards and the advent of new digital diagnostic testing methods.

Several challenges were encountered building the PPP. First, providing private healthcare facilities with access to public COVID-19 testing facilities could only be realized by positioning the PPP as a ‘scientific research project’. This implied developing a full research proposal along the KEMRI format and submission to various entities for ethical clearance. The process was accelerated by applying a 2-step procedure: first going for an expedited operational clearance at the Kisumu County level (1 month) to get COVID-Dx started and next to go in parallel for more extensive clearance at the national level (6 months). Our general recommendation from this experience is to install Ethical Review processes at the national (MoH) level in Kenya that can act within weeks and are allowed to be used when a health emergency is declared by the government.

Second, operational and material challenges for the PPP were frequently encountered, including high costs and lack of adequate laboratory supplies, laboratory equipment repair delays, and procuring PPE supplies for healthcare facilities. The essential PPE supplies were not readily available in Kenya when the project started and when available their prices were grossly overrated. Given the urgency to respond to the pandemic, we started with paying approximately double the price of PPEs. Waiting a few months into the project for prices to fall was not tenable at the time and would have resulted in significant delays. Throughout the project, the supply chain of PPEs was restored, and prices went gradually down, although laboratory equipment breakdown and supply chain disruptions remained a regular challenge. In general, the emergency situation required continuous adaptations and frequent consultations, which were facilitated by PharmAccess NGO. This helped building trust between the stakeholders and facilitated developing task divisions rapidly and naturally.

COVID-19 related stigma appeared much more important than expected. Some private facilities were afraid of being labelled COVID-19 testing sites. This fear was partially the result of (lockdown) measures implemented by government authorities early in the pandemic. When a public facility had staff who tested positive for COVID-19, the authorities closed the facility. Loss of income due to such measures was reportedly an important reason for private facilities to participate in COVID-Dx. To mitigate this, our project concentrated on testing healthcare staff at the participating facilities (and elsewhere), to keep them optimally informed, and in case of positivity, provide mental support services during the quarantines. Additionally, the facilities hesitated to join COVID-Dx because they were afraid patients would avoid visiting them out of fear of being infected, which would also reduce the number of patients with chronic conditions. To mitigate these fears, a letter from the Kisumu DoH was secured in which it was reassured that private facilities would not be closed if any of their patients tested positive for COVID-19. At the onset of COVID-Dx some reference to moral responsibilities of private facilities to pick up their role appeared important. When the first facilities eventually joined, this created confidence for other private facilities to follow suit.

To effectively combat COVID-19 stigma the COVID-Dx project recruited a senior Psychosocial Counsellor to provide direct services to clients. During the roll-out these services appeared very much appreciated and needed scaling. Thus, the senior counsellor trained the existing HIV counsellors of participating providers to also address the mental challenges of COVID-19 (testing). All counsellors underwent a training from the County’s Mental Health and Psychosocial Support department as well as refreshers from the Senior Counsellor. The counsellors remained active throughout the project and were a great help to the facilities, the outreach team, and patients in contact tracing, encouraging contacts to test, reporting results to participants as part of post-test counselling and addressing patient fears around COVID-19 stigma. This intervention helped reducing fear, stigma, depression and other mental health related cases among the patients and health care workers.

During COVID-Dx, decisions by public entities sometimes had consequences for the set research protocols and processes in the project. For instance, the MoH revised the national guidelines on COVID-19 testing several times throughout the project, and some sample taking sites closed unilaterally without consulting other partners. The COVID-Dx project team continuously tried to manage all parties and adhere to National Guidelines. Additionally, the team requested the highest decision-makers in Kisumu County to have some decisions reviewed, based on practical experiences in the field.

Challenges identified by other studies of PPPs in the healthcare sector in LMICs include providing diagnostics, the capacity to train and supervise private providers, disruptions in funding, slow implementation of the public sector, lack of information sharing, and mismatched organizational styles and differing priorities^38-41^. Findings from Ghana implied that NGOs could be valuable to government for their ability to increase reach and to offer technical expertise^39^. A different Kenyan study suggested consistency and flexibility are crucial to make PPPs successful^38^. These findings confirm our PPP experiences. We learned, and literature also confirms, that to prevent, control and manage future outbreaks before these become epidemics, sub-Saharan African countries will need investments and political will so public health resources, PPPs and scientific expertise can be aligned^42^. Additionally, we learned integrating digital technologies markedly improve and supports the responses to a pandemic. Digitalization of the entire patient COVID-19 service trajectory allowed for contraction and expansion whenever needed due to case load of COVID-19 patients. It increased transparency of funding channels, improved quality and efficiency of health service delivery and empowered policy makers to make data-based decisions in semi-real time.

### Quantitative insights

COVID-Dx encountered an overall operational COVID-19 positivity rate of 9.8%. This figure should not be considered as representative for the Kisumu general population, since the number of weekly tests fluctuated due to operational issues, political decisions, or social challenges. Nevertheless, the positivity rate graph served as guidance for the project team to manage the project in terms of operational challenges, like staff fluctuations, procurement of supplies and utensils, transport arrangements. The project proved epidemically prepared, starting with a 0% positivity rate in June 2020 and moving upwards after the first positive cases were reported in Kisumu^43^. The highest operational positivity rate was by December 2020 (23.1%), which is probably related to increased traveling and human contacts due to the festive season, creating the Kenyan ‘second wave’, which was more severe than the first wave^44,45^.

The overall operational positivity rates varied extensively between the participating facilities. One explanation for the high overall positivity rate (31.9%) of Facility H could be that this facility only joined COVID-Dx in December 2020, just in the middle of the second wave. Facility I also showed interesting dynamics, as its first tests were performed in February of 2021, which was also in the period of more COVID-19 cases in the county, yet the positivity rate remained relatively low at 5.6%. This was maybe due to high testing volumes amongst more general population; 662 tests were done in two months. Facility E and F terminated the project in November 2020. COVID-19 politics within the County and ethical concerns in one of the facilities which did not align with the MoH Code of Ethics, were reasons to stop collaboration with these facilities. November 2020 had the lowest positivity rate, mostly due to a temporary suspension of testing at some facilities, due to lack of PCR reagents and supplies at the central laboratories. We noted rising positivity rates at the end of this first COVOD-Dx episode in March 2021 which mirrored the start of the third COVID-19 wave (Delta) in Kisumu County.

The average age of COVID-19 positive patients (40.2 years) appeared significantly higher than negative tested patients (36.4 years) (two-sample t-test (M= 36.8, SD = 15.4), 95% CI [36.3, 37.3], t = -4.8, *P* = 0.00). The data also showed older positive adults (34 and above) reported considerably more clinical symptoms than younger patients. These observations are in line with other COVID-19 epidemiological data in Kenya^46^ and elsewhere^47,48^. Importantly, the socio-demographic profiles of tested patients in COVID-Dx are not representative for Kisumu County or Kenya, since patients were selected according to regularly changing MoH national testing guidelines and priorities, due to lack of diagnostic supplies.

We considered healthcare workers a priority in the project. Goal was to keep them informed, working, and supported, in line with the MoH priorities. The project managed to perform 1,009 tests, equivalent to almost half (43%) of the Kisumu healthcare work force. Testing healthcare workers regularly also ensured they felt safe while executing their work, motivating them to continue sample-taking and assisting patients. At a later stage when vaccinations became available, a disproportional percentage of COVID-19 break-through infections was observed, resulting in an immediate policy advice to booster this target group (results to be published elsewhere).

Developing the CommCare App was a continuous process, with constant improvements to adapt to the needs of the healthcare facilities, needs in sample tracking and transportation, and reporting needs from the laboratories. These changes sometimes affected how we presented the data in the final database. Regular extensive data cleaning was required, which was done manually regularly, the database issues were resolved in time for the final data analyses. It is also vital to highlight that digitization of CCIF was a unique aspect of this project-which promoted quick data transmission compared to paper forms employed by MOH during that time. Health care facilities were motivated to use the COVID-Dx tools, because they could directly see their inputs uploaded into the dashboards visible on their mobile phones.

While first focusing on building a robust PPP model, during COVID-Dx we explored other opportunities to scale up. Early in the project, we experienced the substantial limitations of large-scale PCR testing, such as high costs, requirement of trained staff, sophisticated equipment, and good logistics. Often it was not possible to test all symptomatic patients. Moreover, even if testing was available, the time to report back testing results to patients took long. The original goal of 24-48 hours was often not achieved and certainly not realized during the Christmas holiday season. Prolonged turnaround time for COVID-19 resulted in increased patient anxiety, impaired tracking of cases/contacts and hampered public health efforts against the pandemic. To exacerbate these diagnostic challenges and in the midst of stockouts of COVID-19 PCR test kits, VTMs and PPEs, limited molecular testing capacity seriously hampered efforts to tame to pandemic.

All these challenges led us to explore the possibilities of rapid testing for COVID-19 using antigen testing. The costs of RDTs are lower, results can be available within 20 minutes, no equipment is necessary, and less training is required. A sub-study within COVID-Dx was implemented to validate RDTs and compare them to PCR tests. The favourable results of this sub-study are published in a different paper^33^.

Trust gained through COVID-Dx during the first part of the pandemic, led to a closer working relationship with Kisumu County to fight the next phase of the epidemic: the outbreak of the Delta variant in sub-Saharan Africa^5^. As described in this paper, in May 2021, Kisumu County health officials decided to deploy the existing COVID-Dx network as a County-wide approach. PharmAccess became technical assistance partner to realize a COVID-Dx dashboard for Kisumu. This dashboard became accessible to key County policymakers and decision-makers, showing COVID-19 hotspots, positive cases, vaccination status, etc. Moreover, based on direct requests of DoH and participating health care providers a dashboard was developed that indicated availability of oxygen, numbers of hospital beds, ambulances, diagnostic tests, etc. This largely helped facilities encountering shortages to link up to neighbouring facilities that according to the dashboard still had surpluses. Rapid scalability of COVID-Dx was thus proven on several occasions, with its digitalized trajectory monitoring tools and dashboard playing a crucial role. More detailed data are currently being analysed and described in separate papers. COVID-Dx is therefore a good example in Kenya of PPP epidemic preparedness^49^.

## Conclusion

Sudden health emergencies such as pandemics are a serious challenge to already strained health systems in resource-poor settings, particularly in sub-Saharan Africa. In those circumstances, all available infrastructure and manpower should be deployed. This paper describes a successful PPP, involving both public and private sector health facilities in Western Kenya. We demonstrated feasibility by taking a ‘can do’ approach and addressing operational challenges step-by-step as they unfolded, facilitated by a coordinating NGO. The model was proven scalable in practice, expanding to Kisumu (and very recently even to 13 other Counties) and can serve as an example of PPPs for epidemic preparedness in SSA. Specific additional strength of this approach was the investment in digitalization and digital dashboards for project monitoring, patient service delivery, health facility improvement and rapid data processing to inform policy makers and health managers. This significantly accelerated operational decision-making, like timely identification of COVID-19 hotspots so DoH outreach teams could be deployed efficiently. Building robust epidemic preparedness benefits from African health systems to go digital. Such digital infrastructures are flexible, can contract and expand with (waves of) epidemics and can be rapidly scaled to other geographic areas and communities, as proven by this PPP. The future of health systems strengthening in Africa is going digital.

## Data Availability

Due to the privacy of patient data, datasets associated with this project are not publicly available. Anonymized and aggregated data are available from the authors upon request.

## Abbreviations

CCIF: COVID Case Identification Form
COVAX: COVID-19 Global Vaccine Access
COVID-19: coronavirus disease of 2019
DoH: Department of Health
GDPR: General Data Protection Regulation
GFATM: The Global Fund to Fight
AIDS: Tuberculosis and Malaria
HIPAA: Health Insurance Portability and Accountability Act of 1996
KEMRI: Kenya Medical Research Institute –
LMIC: low- and middle-income countries
MoH: Ministry of Health
NGO: non-governmental organization
PCR: polymerase chain reaction
PEPFAR: President’s Emergency Plan for AIDS Relief
PPE: personal protective equipment
PPP: public-private partnership
RDT: rapid diagnostic test –
RNA: Ribonucleic acid
ToT: trainers of trainers
UHC: universal health coverage
VTM: viral transport medium –
WHO: World Health Organization

## Acknowledgements

The authors would like to express gratitude to all clinicians, nurses, lab technicians, motorbike riders, data entry personnel and all other personnel who were active in the field during this study. The CommCare mobile data collection software was provided by Dimagi free of charge. We would also like to thank the Kisumu Department of Health for its guidance and cooperation to execute this project. We would like to thank our funders: Achmea Foundation, Pfizer Foundation, and the Netherlands Ministry of Foreign Affairs.

## Contributor-ship statement

TRW designed the project and the research study, and additionally scientifically supervised the project. NH assisted with the design and implementation of the study and managed the operations in Kenya of this project. SK and AO oversaw research activities and were involved at decision-making level. HB contributed to the project design and implementation and managed KEMRI’s role. EM was responsible for onsite day-to-day operations during the project and managed relationships between all partners. MO assisted in the field and was responsible for the social sciences, with assistance from IO. RA managed our psychosocial support on the ground. AK regularly visited facilities for data management and managed the database. TS developed and updated the dashboard. MJ supported with clinical supervision. KO managed the laboratory aspects of this study. SO contributed research, such as the RDT evaluation. CW managed relations with our donors and reported back results to them. SD conducted data analyses, wrote the manuscript, and submitted this manuscript.

## Ethics approval

Ethical clearance for this project was obtained from Jaramogi Oginga Odinga Teaching & Referral Hospital (JOOTRH) on June 16, 2020, with approval number IERC/JOOTRH/230/2020. KEMRI also provided ethical clearance on September 30, 2020, with approval number KEMRI KEMRI/SERU/CGHR/05-05/4038. Research License was obtained from the National Commission for Science, Technology, and Innovation (NACOSTI) on July 6, 2020 (NACOSTI/P/20/5616). The roll-out of this project was in complete coordination with the Kisumu Department of Health. Sample collection in patients in this project was voluntary.

## Contribution to the field

The findings of this study provide a practical contribution how African health system epidemic preparedness can rapidly scale through PPP and digitalization. The ‘learning while doing’ approach that was chosen in view of the urgency of the COVID-19 outbreak proved effective and scalable. This was further strengthened by the coordination of a local NGO, PharmAccess, keeping communication lines open, performing administrative functions as required by the health system and rapidly securing complementary funding to address important developments during the various COVID-19 waves in Kisumu, Kenya. In addition, this PPP was unique as it was strongly supported by integrated digital technologies on mobile phones of all key players, allowing for data-based policy decision making. The digital data entry App with corresponding dashboards that were co-designed according to local needs ensured transparency, efficiency and frequent usage. All in all, ‘digital’ is the way going forward strengthening African health systems, with the concomitant advantage of speed and flexibility when combatting sudden epidemic outbreaks and the potential for rapid scaling into regional interventions.

