## Supplementary materials for "Public-private partnership to rapidly strengthen and scale COVID-19 response in Western Kenya"

**Figure 1. Screenshots Operational Dashboard COVID-Dx**

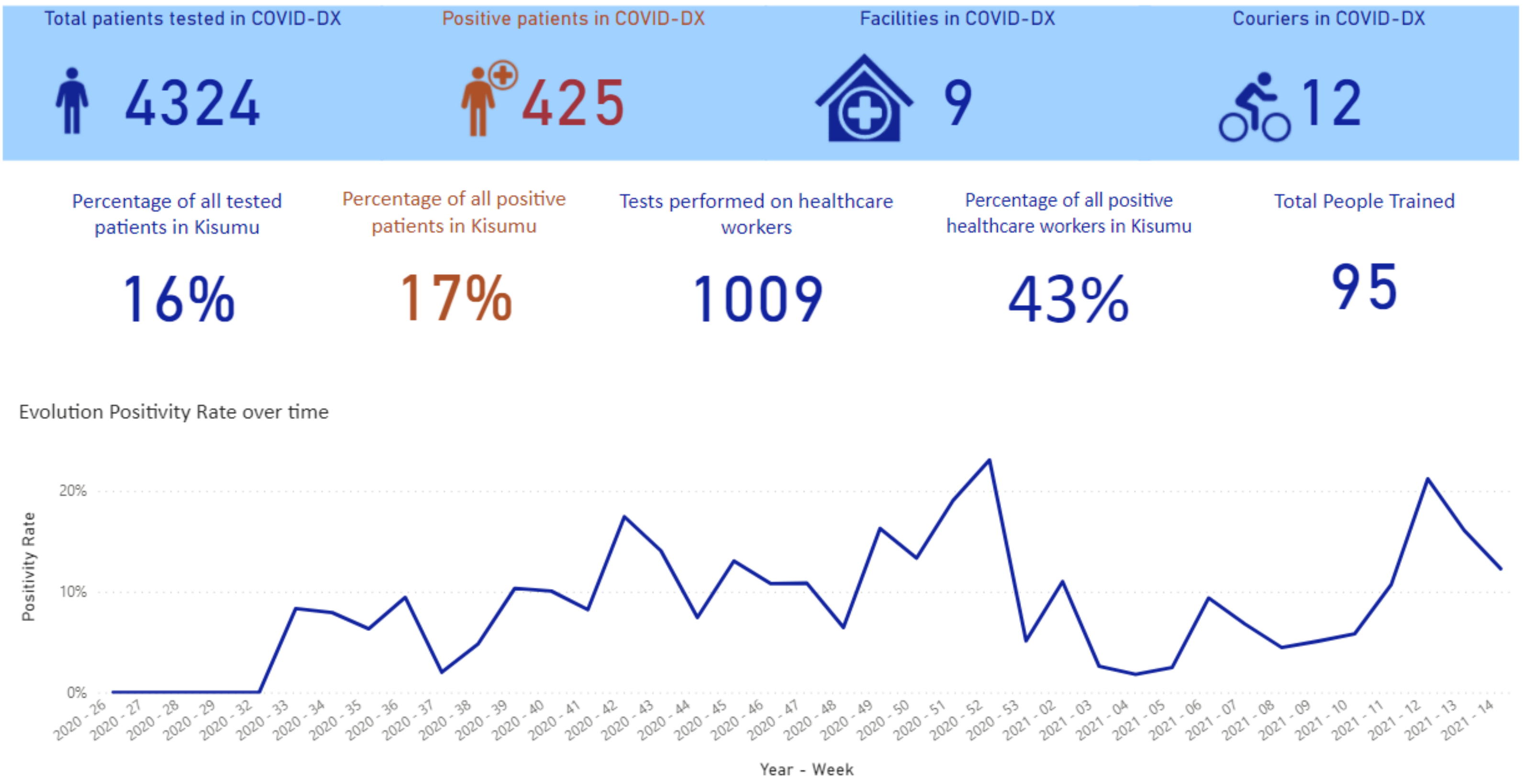

##### Testing results

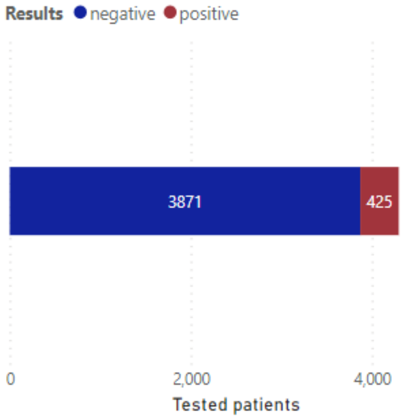


*Total patients tested grouped by results.*

##### Patients enrolled in COVID-DX per day

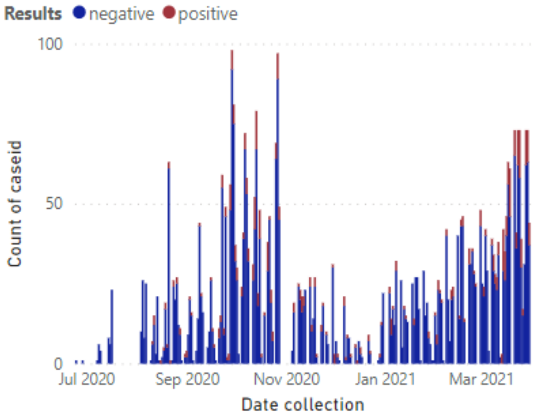


*Overview of the total number of patients enrolled over time.*

##### Time between sample collection and data entry completion

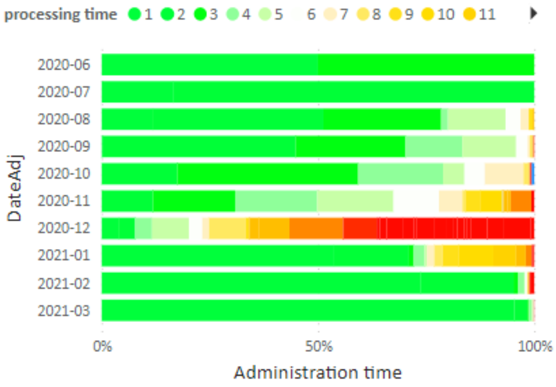


*Number of days between sample collection and results reporting*

##### Positive patients reported asymptomatic

# 23%

##### Frequency with which symptoms have been reported in positive cases

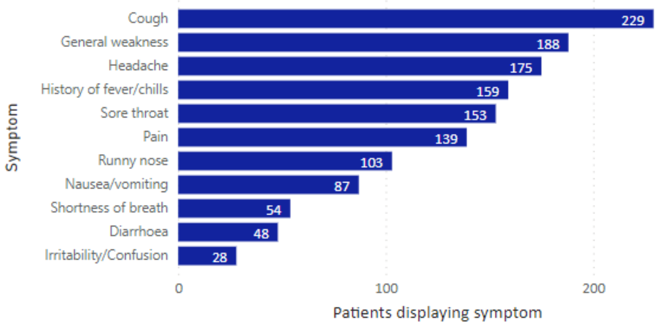


*In the graph above, a patient can be represented multiple times if (s)he displayed multiple symptoms.*

##### Underlying conditions (when applicable)

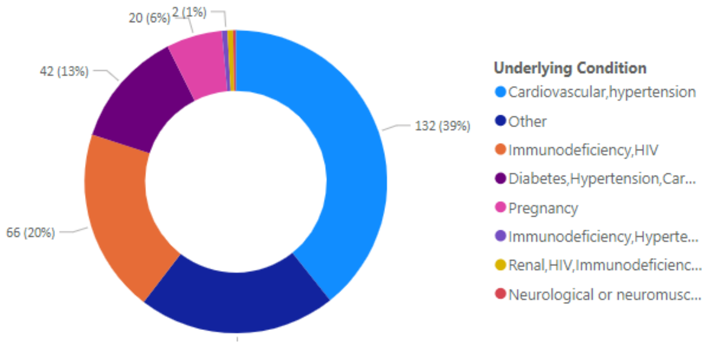


Supplementary Materials Manuscript "Public-private partnership to strengthening COVID-19 response in Kisumu, Kenya"

### Figure 2. COVID-Dx roll-out in the context of Kenyan COVID pandemic highlights

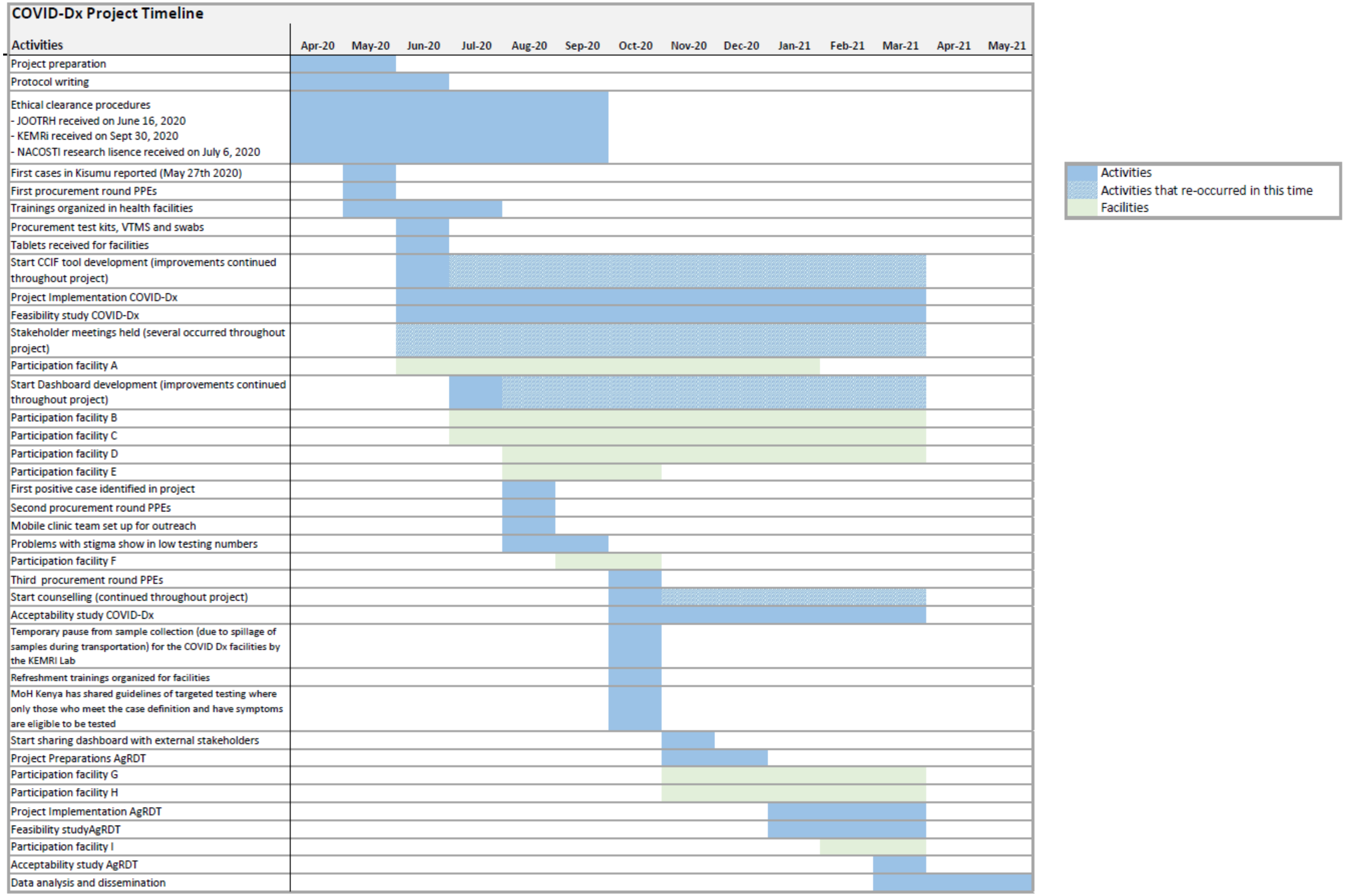
